# SARS-CoV-2/COVID-19 hospitalised patients in Switzerland: a prospective cohort profile

**DOI:** 10.1101/2020.12.10.20246884

**Authors:** Amaury Thiabaud, Anne Iten, Carlo Balmelli, Laurence Senn, Nicolas Troillet, Andreas Widmer, Domenica Flury, Peter W. Schreiber, Miriam Vázquez, Lauro Damonti, Michael Buettcher, Danielle Vuichard-Gysin, Christoph Kuhm, Alexia Cusini, Thomas Riedel, Yvonne Nussbaumer, Roman Gaudenz, Ulrich Heininger, Christoph Berger, Franziska Zucol, Sara Bernhard-Stirnemann, Natascia Corti, Petra Zimmermann, Anita Uka, Anita Niederer-Loher, Céline Gardiol, Maroussia Roelens, Olivia Keiser

**Affiliations:** Institut de Santé Globale, Faculté de Médecine de l’Université de Genève, Geneva, Switzerland; Service de prévention et contrôle de l’infection, Direction médicale et qualité, HUG, Geneva, Switzerland; Infection Control Programme, EOC Hospitals, Ticino, Switzerland; Service de médecine préventive hospitalière, CHUV, Lausanne, Switzerland; Service of Infectious Diseases, Central Institute, Valais Hospitals, Sion, Switzerland; Department of Infectious Diseases, University Hospital Basel, Basel, Switzerland; Division of Infectious Diseases and Hospital Epidemiology, Cantonal Hospital St. Gallen, St. Gallen, Switzerland; Division of Infectious Diseases and Hospital Epidemiology, University Hospital Zurich and University of Zurich, Zurich, Switzerland; Department of Infectious Diseases, Bern University Hospital (Inselspital), Bern, Switzerland; Paediatric Infectious Diseases, Department of Paediatrics, Children’s Hospital, Cantonal Hospital Lucerne, Switzerland; Department of Infectious Diseases, Thurgau Cantonal Hospital, Thurgau, Switzerland; Department of Infectious Diseases, Cantonal Hospital Graubuenden, Chur, Switzerland; Department of Pediatrics, Cantonal Hospital Graubuenden, Chur, Switzerland; Klinik für Innere Medizin, Kantonsspital Spitäler Schaffhausen, Schaffhausen, Switzerland; Innere Medizin und Infektiologie, Kantonsspital Nidwalden, Stans, Switzerland; Infectious Diseases and Vaccinology, University of Basel Children’s Hospital, Basel, Switzerland; Division of Infectious Diseases, and Children’s Research Center, University Children’s Hospital Zurich, Zurich, Switzerland; Paediatric Infectious Diseases, Department of Paediatrics, Cantonal Hospital Winterthur, Winterthur, Switzerland; Children’s Hospital Aarau, Aarau, Switzerland; Unit of General Internal Medicine, Hirslanden Clinic, Zurich, Switzerland; Faculty of Science and Medicine, University of Fribourg, Fribourg, Switzerland; Department of Paediatrics, Fribourg Hospital HFR, Fribourg, Switzerland; Children’s Hospital of Eastern Switzerland, St. Gallen, Switzerland; Swiss Federal Office of Public Health, Bern, Switzerland

## Abstract

**Background:** SARS-CoV-2/COVID-19, which emerged in China in late 2019, rapidly spread across the world causing several million victims in 213 countries. Switzerland was severely hit by the virus, with 43’000 confirmed cases as of September 1st, 2020.

**Aim:** In cooperation with the Federal Office of Public Health, we set up a surveillance database in February 2020 to monitor hospitalised patients with COVID-19 in addition to their mandatory reporting system.

**Methods:** Patients hospitalised for more than 24 hours with a positive PCR test, from 20 Swiss hospitals, are included. Data collection follows a custom Case Report Form based on WHO recommendations and adapted to local needs. Nosocomial infections were defined as infections for which the onset of symptoms started more than 5 days after the patient’s admission date.

**Results:** As of September 1st, 2020, 3645 patients were included. Most patients were male (2168 - 59.5%),and aged between 50 and 89 years (2778 - 76.2%), with a median age of 68 (IQR 54-79). Community infections dominated with 3249 (89.0%) reports. Comorbidities were frequently reported: hypertension (1481 - 61.7%), cardiovascular diseases (948 - 39.5%), and diabetes (660 - 27.5%) being the most frequent in adults; respiratory diseases and asthma (4 −21.1%), haematological and oncological diseases (3 – 15.8%) being the most frequent in children. Complications occurred in 2679 (73.4%) episodes, mostly for respiratory diseases (2470 - 93.2% in adults, 16 – 55.2% in children), renal (681 – 25.7%) and cardiac (631 – 23.8%) complication for adults. The second and third most frequent complications in children affected the digestive system and the liver (7 - 24.1%). A targeted treatment was given in 1299 (35.6%) episodes, mostly with hydroxychloroquine (989 - 76.1%). Intensive care units stays were reported in 578 (15.8%) episodes. 527 (14.5%) deaths were registered, all among adults.

**Conclusion:** The surveillance system has been successfully initiated and provides a very representative set of data for Switzerland. We therefore consider it to be a valuable addition to the existing mandatory reporting, providing more precise information on the epidemiology, risk factors, and clinical course of these cases.

## Introduction

With almost 27 million confirmed infections, of which 70.5% recovered, and more than 850’000 deaths [1], the Severe Acute Respiratory Syndrome Coronavirus 2 (SARS-CoV-2)/ Coronavirus disease 2019 (COVID-19) pandemic has had a massive impact on all aspects of everyday life; including politics, travels, and economics. The virus was initially identified in a cluster of Chinese adult patients with pneumonia of unknown cause [2] incriminating a large seafood and animal market in Wuhan City as the source of the outbreak; but rapidly spread via person-to-person transmission. Since then, 213 countries and territories have been affected [3].

On January 30th 2020, the World Health Organisation (WHO) declared the outbreak a Public Health Emergency of International Concern (PHEIC). WHO emphasized the urgent need to coordinate international efforts to investigate and better understand this novel coronavirus, to minimize the threat in affected countries and to reduce the risk of further international spread [4]. WHO designed various study protocols for early investigation of the outbreak, including a global Anonymized Clinical Data Platform (the “nCoV Data Platform”) to enable State Parties of the International Health Regulations (IHR) (2005) to share with WHO anonymized clinical data and information related to patients with suspected or confirmed infections with the disease (collectively “Anonymized nCoV Data”) [5].

The first patient with COVID-19 in Switzerland was reported on February 25th, 2020, in the canton of Ticino, earlier than in most European countries due to its proximity and cross-border commuting with Italy. In order to prevent and monitor the spread of the disease in Switzerland, suspicion of COVID-19 cases by clinicians were reported to the Federal Office of Public Health (hereafter FOPH) through an obligatory declaration form [6]. The number of confirmed cases rose rapidly to more than 8000 positive cases and 66 deaths within only a month after the start of the epidemic in Switzerland. Compared to its neighbour countries, Switzerland has similarly suffered from the pandemic with a cumulative incidence of 496.5 confirmed cases per 100’000 habitants (Germany - 264.7, France - 449.9, Italy - 448.4) but with an overall lower cumulative death incidence of 200.9 deaths per 1’000’000 inhabitants (Germany - 111.6, France - 471.2, Italy - 586.2) as of September 4th, 2020 [7].

Early analysis and preparations for the advent of the COVID-19 pandemic in Switzerland by officials, including identification of risk groups and forecasting, were based on Chinese data. However, the Chinese population differs from the Swiss population, and such models were therefore misleading and inaccurate. Based on our experience with the hospital-based surveillance of influenza cases in Switzerland [8] and in collaboration with the FOPH, we developed a surveillance system of hospitalised COVID-19 cases in early February. We aim to:

1. Test the adaptability of the influenza system to other infectious diseases and quantify the workload to be able to react faster to future epidemics.
2. Monitor the evolution of hospitalised COVID-19 cases in Switzerland, alongside the mandatory reporting system.
3. Provide a reliable, harmonized, in-depth dataset for hospitalised COVID-19 cases in Switzerland that can be used for decision-making and public health purposes.

The aim of the current manuscript is to present a profile of patients hospitalised with COVID-19 during the first epidemic wave in Switzerland.

## Study design and procedures

### Inclusion

The study was designed to capture most of the severe COVID-19 cases in Switzerland by including hospitalised patients from 20 large cantonal and university hospitals. To ease the process and to minimize the time of developing the system, we first contacted the participating hospitals of the Influenza pilot surveillance system [8]; seven of which agreed to participate (Hôpitaux Universitaires de Genève, Kantonsspital St Gallen, Ospedaliero Cantonale Ticino, Hôpital du Valais, Centre Hospitalier Universitaire du Canton de Vaud, Universitätsspital Zürich, Universitäts-Kinderspital Zürich). Since patients were more likely to be sent to large hospitals with more capacity and means to treat a novel disease, the system also captures patients from other large university and cantonal hospitals which agreed to participate: Universitätsspital Basel, Inselspital Bern, Kantonsspital Graubünden, Luzerner Kantonsspital (including Kinderspital Luzern), Spitäler Schaffhausen Kantonsspital, Thurgau Hospital Group, and smaller cantonal hospitals (Hôpital de Fribourg, Kantonsspital Aarau, Kantonsspital Winterthur, Kantonsspital Nidwalden, and Klinik Hirslanden Zürich). Three of the smaller hospitals (Fribourg, Aarau, and Winterthur) only contributed with paediatric cases. We also included three paediatric hospitals, separate from their adult counterparts: Universität Kinderspital Basel, Ostschweizer Kinderspital, and Kinderspital Zürich. The Kantonsspital St. Gallen also included cases from all public hospitals of the canton: they were included by the identical infection control team.

Patients, regardless of age or gender, with a polymerase chain reaction (PCR) confirmed COVID-19 diagnosis who were hospitalised for more than 24 hours, were included in the database. The data were collected by infection control specialists or other physicians, dedicated study nurses, or medical students of the participating centres under the supervision of physicians and stored anonymously in a secure REDCap database [9] located in the Clinical Research Center of the University Hospitals of Geneva. Data entry started on March 1^st^, 2020 and is still ongoing. Data were also collected retrospectively.

### Questionnaire

Data are collected based on the Case Report Form (CRF) developed for the Influenza surveillance pilot [8] with relevant COVID-19 additions. This CRF is split in two parts: the compulsory part contains inclusion criteria, demographics, case declaration (classification, date of detection and date of symptoms, laboratory sample information), admission details (hospitalisation ward, origin of patient, severity at admission), and follow-up forms. The optional part describes the patient’s stay in more detail with description of complications, intensive/intermediate care stays, treatment, and risk factors in a clinical complementary information form. The CRF is added as supplementary material.

Since the system was put in place when movement and contact tracing became difficult, we only selected part of the recommended WHO COVID-19 CRF [10] and of the FOPH obligatory declaration form [6]. We therefore removed any information about travels to foreign countries or ethnicity. We, however, kept employment in healthcare facilities as an important risk factor. The follow-up and daily forms of the WHO CRF were deemed too time consuming to retrieve by participating hospitals, and were therefore discarded in favour of the influenza CRF structure. Complications and underlying medical conditions (UMC) were also similar to the Influenza CRF with small additions to conditions shown to be of importance either as risk factors (e.g. hypertension, angiotensin-converting-enzyme (ACE) inhibitor treatments, obesity), or as consequences of the SARS-CoV-2 infection (acute respiratory distress syndrome (ARDS), thrombosis/embolism). To assess the severity of the disease at admission, we also added the CURB-65 score [9] for adults, and a similar severity score for children based on the expertise of the participating paediatric infectious disease specialists. Finally, we classified cases as community-acquired or nosocomial, by defining nosocomial cases to be cases for which the onset of symptoms started more than 5 days after the patient’s admission date. This time limit was to account for the incubation time for the virus.

As a surveillance tool and because of the rapid growth of the epidemic, basic epidemiological information had to be reported within 48h, comprising at minimum the inclusion criteria, demographics, and case declaration forms in order to report the new cases and their location with little delay. Additional information on the hospitalisation itself (admission, follow-up, and clinical information) were filled as soon as possible depending on the severity of the epidemic and local issues.

The database records entries as COVID-19 episodes rather than per patient, with the possibility to link two episodes. A new episode was defined if a patient was readmitted into hospital more than 30 days after their previous discharge. If a transfer between hospitals occurred, a new record was created in the destination hospital. The database and the CRF implementation were tested by the participating sites at the time of deployment and tweaked following their feedback. Small additional changes were made as the database was already in production when new information about the disease became available (e.g. thrombosis/embolism).

The data presented in this manuscript were extracted on August 31st, 2020. The week used in the figures of this manuscript is the week of interest for the system, defined as the week of the sample collection date (equivalent to the diagnosis) for nosocomial cases or the week of admission into the hospital for other cases.

## Characteristics of patients

### Demographics

As of August 31st, 2020, 3650 episodes in 3645 patients have been recorded in the database, with 66 patients having been hospitalised more than once for the same episode. Figure 1 shows the age and gender distribution of patients in the database and Table 1 shows their baseline characteristics. Overall, 2168 (59.5%) patients were male and 1477 (40.5%) female, and the median age was 68 years (IQR 54-79). Most were aged between 50 and 89 years (2778 – 76.2%), followed by patients between 40 and 49 years (299 – 8.2%), patients between 90 and 99 years (215 – 5.9%), and patients between 30 and 39 years (185 −5.1%). Among children, patients between 13 and 19 years and patients between a month and less than a year are dominant, with 24 (0.7%; 35.3% of all children) and 23 (0.6%; 33.8% of all children) patients respectively.

**Table 1.**
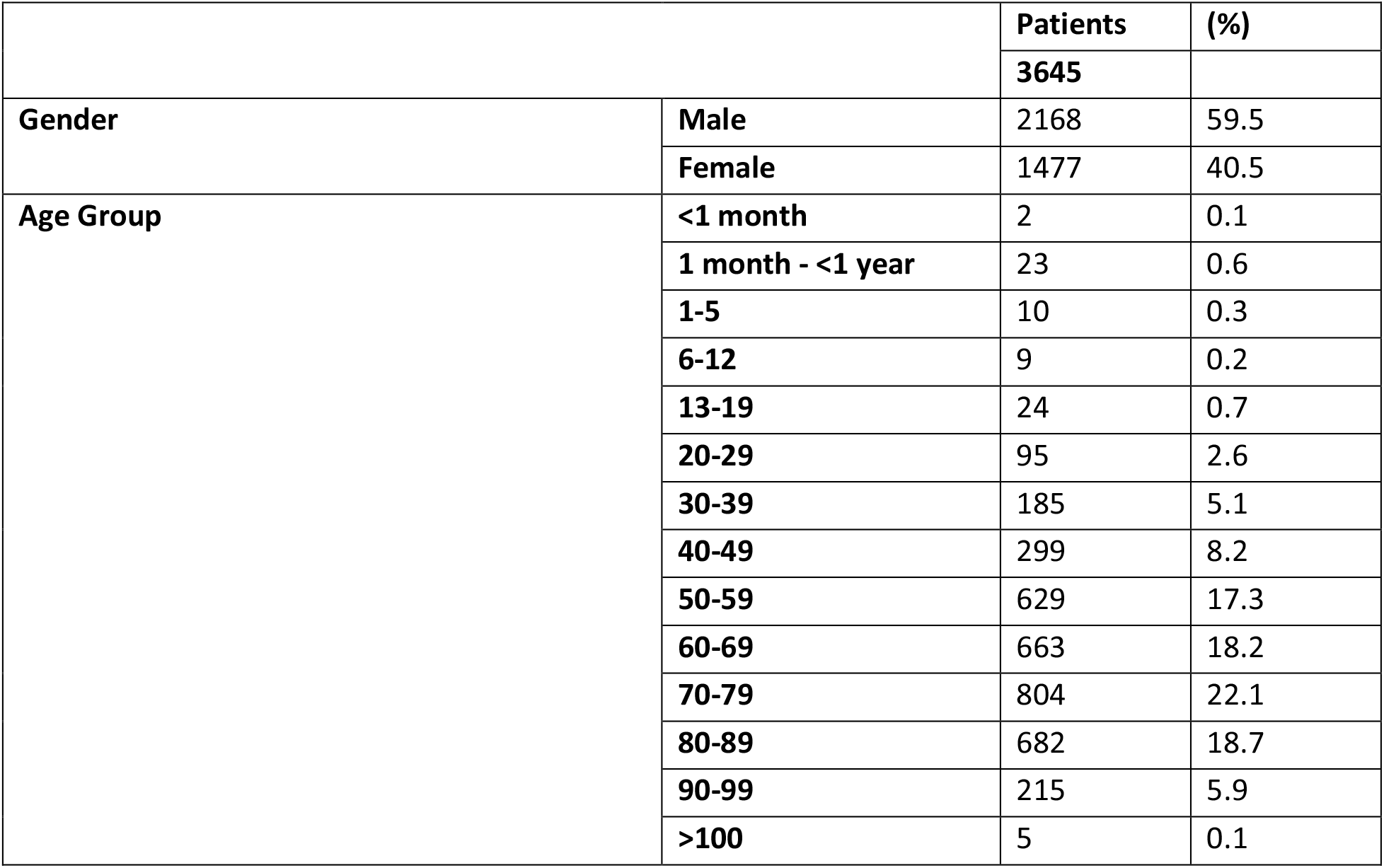
Baseline characteristics of patients registered in the database.

|  |  | Patients | (%) |
| --- | --- | --- | --- |
|  |  | 3645 |  |
| Gender | Male | 2168 | 59.5 |
|  | Female | 1477 | 40.5 |
| Age Group | <1 month | 2 | 0.1 |
|  | 1 month - <1 year | 23 | 0.6 |
|  | 1-5 | 10 | 0.3 |
|  | 6-12 | 9 | 0.2 |
|  | 13-19 | 24 | 0.7 |
|  | 20-29 | 95 | 2.6 |
|  | 30-39 | 185 | 5.1 |
|  | 40-49 | 299 | 8.2 |
|  | 50-59 | 629 | 17.3 |
|  | 60-69 | 663 | 18.2 |
|  | 70-79 | 804 | 22.1 |
|  | 80-89 | 682 | 18.7 |
|  | 90-99 | 215 | 5.9 |
|  | >100 | 5 | 0.1 |

**Figure 1.**
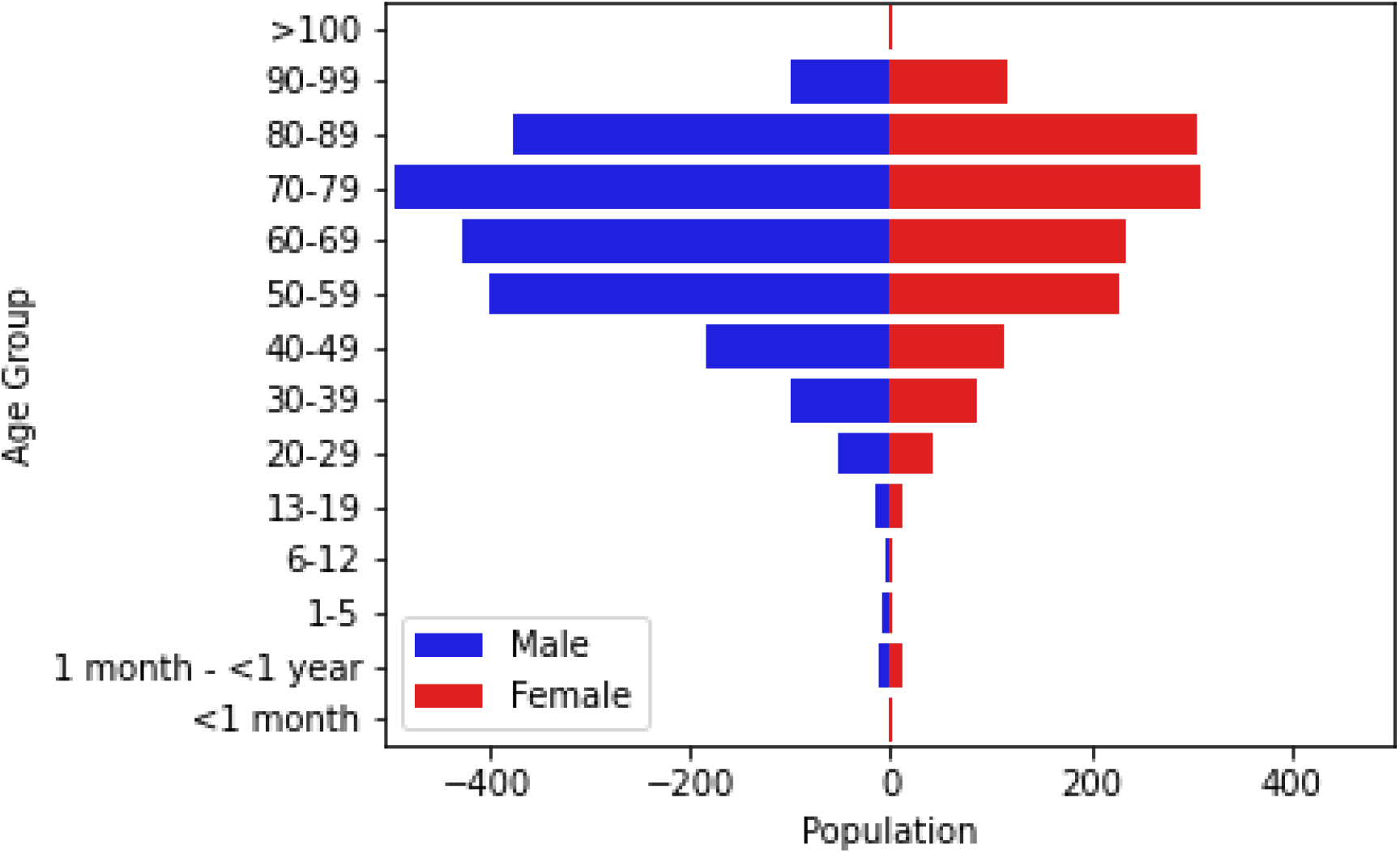
Population breakdown of the 3645 patients hospitalized with COVID 19 between March 1st and August 31st, 2020, by age and gender.

The majority of episodes were community acquired (3249 - 89.0%) while 349 (9.6%) were nosocomial acquired or of unknown source (52 - 1.4%). Nosocomial cases were found less often in children (4.4% of all episodes in children versus 9.7% of all episodes in adults). Figure 2 shows the evolution in the number of episodes throughout the epidemic. The first episodes were recorded in week 2020-09, followed by a rapid rise to about 900 episodes in week 2020-13 before decreasing and stagnating from week 2020-20 onwards to a few episodes declared. A small increase in week 2020-25 was observed. At the time of writing, the number of new cases stabilised to around 30 per week.

**Figure 2.**
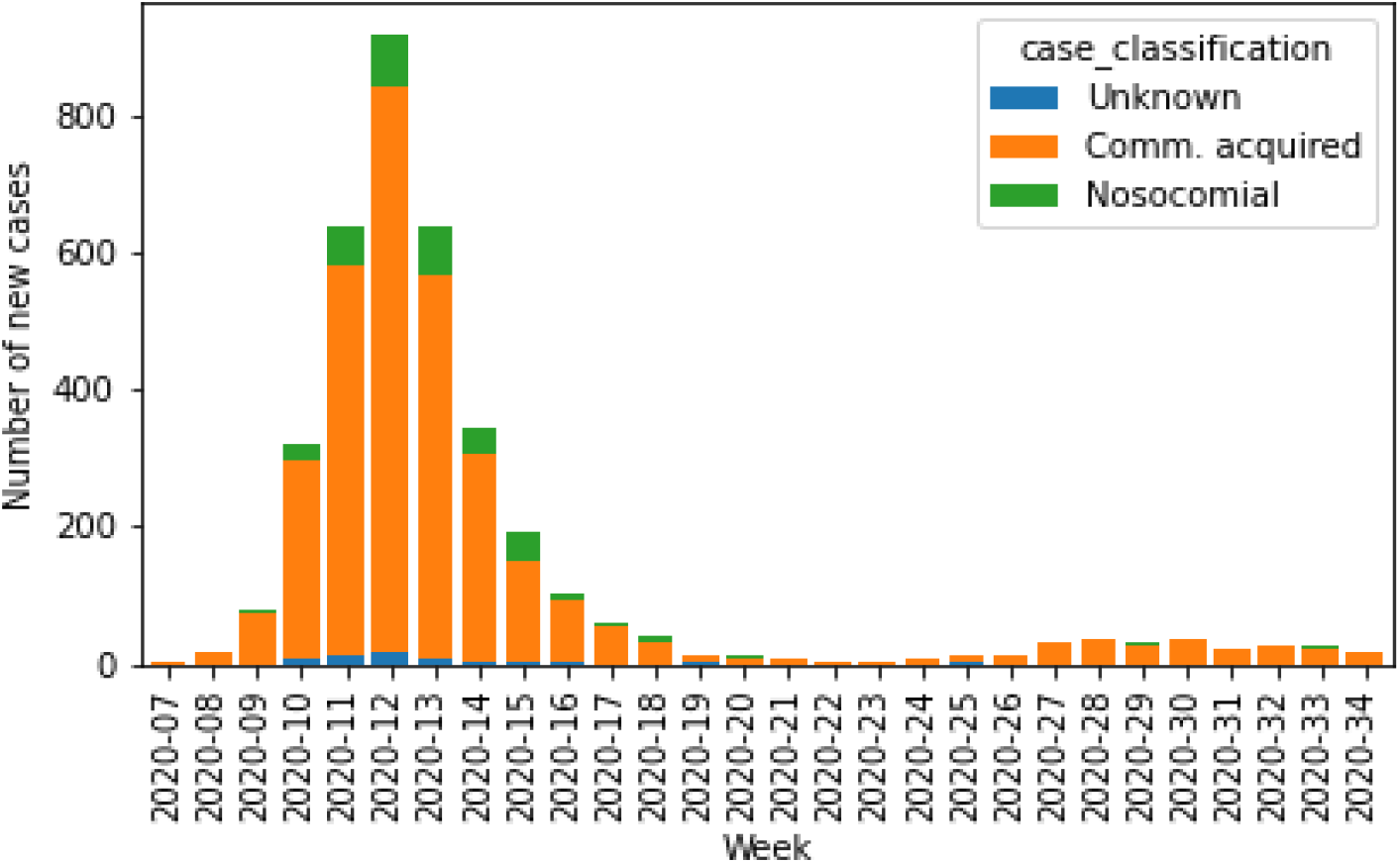
Distribution of SARS-CoV-2/COVID-19 episodes registered in the database by origin of infection.

### Potential risk factors for COVID-19

The risk factors recorded, categorised into comorbidities (typically, chronic diseases), pregnancy status for women, and history of smoking are split by age group for adults and children in Table 2 and 3 respectively. The Charlson Index [12] was used to calculate an overall risk score which median value was 3.0 (IQR 1-6) for both the overall and adult patients. This score had a median value of 0.5 (IQR 0-1.75) among children.

**Table 2.** Characteristics of SARS-CoV-2/COVID-19 episodes, outcomes, and related consequences (complications) encountered in registered episodes for adults (age groups 20-29 to >100).

|  |  | Episodes | (%) |
| --- | --- | --- | --- |
|  |  | 3582 |  |
| Case Classification | Unknown | 47 | 1.3 |
|  | Community acquired | 3189 | 89 |
|  | Nosocomial | 346 | 9.7 |
| Smoked in the last 10 years | Unknown | 1537 | 42.9 |
|  | No | 1808 | 50.5 |
|  | Yes | 237 | 6.6 |
| Pregnancy - Female between 15 & 50 only | Unknown | 381 | 65.8 |
|  | No | 163 | 28.2 |
|  | Yes | 35 | 6 |
| Had at least one comorbidity | Unknown | 574 | 16 |
|  | Yes | 2401 | 67 |
|  | No | 607 | 16.9 |
| ...Respiratory diseases | Unknown | 3 | 0.1 |
|  | No | 1883 | 78.4 |
|  | Yes | 515 | 21.4 |
| ...Asthma | Unknown | 4 | 0.2 |
|  | No | 2193 | 91.3 |
|  | Yes | 204 | 8.5 |
| ...Diabetes | Unknown | 1 | 0 |
|  | No | 1740 | 72.5 |
|  | Yes | 660 | 27.5 |
| ...Hypertension | Unknown | 3 | 0.1 |
|  | No | 917 | 38.2 |
|  | Yes | 1481 | 61.7 |
| ...Cardiovascular diseases | Unknown | 4 | 0.2 |
|  | No | 1449 | 60.3 |
|  | Yes | 948 | 39.5 |
| ...Renal diseases | Unknown | 4 | 0.2 |
|  | No | 1897 | 79 |
|  | Yes | 500 | 20.8 |
| ...Liver diseases | Unknown | 3 | 0.1 |
|  | No | 2254 | 93.9 |
|  | Yes | 144 | 6 |
| ...Neurological impairment | Unknown | 8 | 0.3 |
|  | No | 2007 | 83.6 |
|  | Yes | 386 | 16.1 |
| ...Haematological diseases with immunosuppression | Unknown | 2 | 0.1 |
|  | No | 2342 | 97.5 |
|  | Yes | 57 | 2.4 |
| ...Oncological diseases | Unknown | 7 | 0.3 |
|  | No | 2079 | 86.6 |
|  | Yes | 315 | 13.1 |
| ...Rheumatological diseases with immunosuppression | Unknown | 2 | 0.1 |
|  | No | 2321 | 96.7 |
|  | Yes | 78 | 3.2 |
| ...Dementia | Unknown | 12 | 0.5 |
|  | No | 2191 | 91.3 |
|  | Yes | 198 | 8.2 |
| ...Transplantation of solid organs | Unknown | 4 | 0.2 |
|  | No | 2374 | 98.9 |
|  | Yes | 23 | 1 |
| ...HIV positive | Unknown | 45 | 1.9 |
|  | No | 2341 | 97.5 |
|  | Yes | 15 | 0.6 |
| ...Under immunosuppressive treatment | Unknown | 4 | 0.2 |
|  | No | 2284 | 95.1 |
|  | Yes | 113 | 4.7 |
| ...Tuberculosis | Unknown | 16 | 0.7 |
|  | No | 2363 | 98.4 |
|  | Yes | 22 | 0.9 |
| ...Other diseases | Unknown | 3 | 0.1 |
|  | No | 968 | 40.3 |
|  | Yes | 1430 | 59.6 |
| Had at least one complication | Unknown | 587 | 16.4 |
|  | No | 345 | 9.6 |
|  | Yes | 2650 | 74 |
| ... Ear/Nose/Throat complications | Unknown | 18 | 0.7 |
|  | No | 2439 | 92 |
|  | Yes | 193 | 7.3 |
| ...Acute Otitis Media (among ENT) | Unknown | 3 | 1.6 |
|  | No | 182 | 94.3 |
|  | Yes | 8 | 4.1 |
| ...Respiratory complications | Unknown | 6 | 0.2 |
|  | No | 174 | 6.6 |
|  | Yes | 2470 | 93.2 |
| ...Acute Respiratory Distress Syndrome (ARDS - among resp.) | Unknown | 10 | 0.4 |
|  | No | 1903 | 77 |
|  | Yes | 557 | 22.5 |
| ...Pneumonia (among resp.) | Unknown | 19 | 0.8 |
|  | No | 289 | 11.7 |
|  | Yes | 2162 | 87.5 |
| ...Pneumonia linked to COVID-19 (among pneumonia) | Unknown | 9 | 0.5 |
|  | No | 57 | 2.6 |
|  | Yes | 2096 | 96.9 |
| ...Cardiovascular complications | Unknown | 32 | 1.2 |
|  | No | 1987 | 75 |
|  | Yes | 631 | 23.8 |
| ...Digestive complications | Unknown | 41 | 1.5 |
|  | No | 2111 | 79.7 |
|  | Yes | 498 | 18.8 |
| ...Liver complications | Unknown | 28 | 1.1 |
|  | No | 2112 | 79.7 |
|  | Yes | 510 | 19.2 |
| ...Renal complications | Unknown | 28 | 1.1 |
|  | No | 1941 | 73.2 |
|  | Yes | 681 | 25.7 |
| ...Neurological impairment complications | Unknown | 44 | 1.7 |
|  | No | 2170 | 81.9 |
|  | Yes | 436 | 16.5 |
| ...Osteo-articular complications | Unknown | 34 | 1.3 |
|  | No | 2489 | 93.9 |
|  | Yes | 127 | 4.8 |
| ...Thrombosis/Embolism | Unknown | 34 | 1.3 |
|  | No | 2406 | 90.8 |
|  | Yes | 210 | 7.9 |
| ...Other bacterial infection | Unknown | 38 | 1.4 |
|  | No | 2283 | 86.2 |
|  | Yes | 329 | 12.4 |
| ...Other non-bacterial infection | Unknown | 33 | 1.2 |
|  | No | 2485 | 93.8 |
|  | Yes | 132 | 5 |
| ...Other complications | Unknown | 19 | 0.7 |
|  | No | 1475 | 55.7 |
|  | Yes | 1156 | 43.6 |
| Stayed in ICU at least once | Unknown | 604 | 16.9 |
|  | No | 2406 | 67.2 |
|  | Yes | 572 | 16 |
| Was treated against the COVID-19 infection | Unknown | 611 | 17.1 |
|  | No | 1676 | 46.8 |
|  | Yes | 1295 | 36.2 |
| ...Hydroxychloroquine | No | 310 | 23.9 |
|  | Yes | 985 | 76.1 |
| ...Interferon | No | 1293 | 99.8 |
|  | Yes | 2 | 0.2 |
| ...Lopinavir/Ritonavir | No | 620 | 47.9 |
|  | Yes | 675 | 52.1 |
| ...Remdesivir | No | 1150 | 88.8 |
|  | Yes | 145 | 11.2 |
| ...Tenofovir | No | 1294 | 99.9 |
|  | Yes | 1 | 0.1 |
| ...Ribavirin | No | 1295 | 100 |
|  | Yes | 0 | 0 |
| ...Other treatment | No | 1126 | 86.9 |
|  | Yes | 169 | 13.1 |
| Outcome | Deceased | 527 | 14.7 |
|  | Discharged and alive | 2923 | 81.6 |
|  | Still hospitalised | 132 | 3.7 |

**Table 3.** Characteristics of SARS-CoV-2/COVID-19 episodes, outcomes, and related consequences (complications) encountered in registered episodes for children and teenagers (age groups <1 month to 13-19).

|  |  | Episodes | (%) |
| --- | --- | --- | --- |
|  |  | 68 |  |
| Case Classification | Unknown | 5 | 7.4 |
|  | Community acquired | 60 | 88.2 |
|  | Nosocomial | 3 | 4.4 |
| Smoked in the last 10 years | Unknown | 44 | 64.7 |
|  | No | 22 | 32.4 |
|  | Yes | 2 | 2.9 |
| Pregnancy - Female between 15 & 50 only | Unknown | 11 | 64.7 |
|  | No | 6 | 35.3 |
|  | Yes | 0 | 0 |
| Had at least one comorbidity | Unknown | 7 | 10.3 |
|  | Yes | 19 | 27.9 |
|  | No | 42 | 61.8 |
| ...Respiratory diseases | No | 15 | 78.9 |
|  | Yes | 4 | 21.1 |
| ...Asthma | No | 15 | 78.9 |
|  | Yes | 4 | 21.1 |
| ...Diabetes | No | 18 | 94.7 |
|  | Yes | 1 | 5.3 |
| ...Hypertension | No | 18 | 94.7 |
|  | Yes | 1 | 5.3 |
| ...Cardiovascular diseases | No | 18 | 94.7 |
|  | Yes | 1 | 5.3 |
| ...Renal diseases | No | 17 | 89.5 |
|  | Yes | 2 | 10.5 |
| ...Liver diseases | No | 17 | 89.5 |
|  | Yes | 2 | 10.5 |
| ...Neurological impairment | No | 17 | 89.5 |
|  | Yes | 2 | 10.5 |
| ...Haematological diseases with immunosuppression | No | 16 | 84.2 |
|  | Yes | 3 | 15.8 |
| ...Oncological diseases | No | 16 | 84.2 |
|  | Yes | 3 | 15.8 |
| ...Rheumatological diseases with immunosuppression | No | 19 | 100 |
|  | Yes | 0 | 0 |
| ...Dementia | No | 19 | 100 |
|  | Yes | 0 | 0 |
| ...Transplantation of solid organs | No | 19 | 100 |
|  | Yes | 0 | 0 |
| ...HIV positive | No | 19 | 100 |
|  | Yes | 0 | 0 |
| ...Under immunosuppressive treatment | No | 17 | 89.5 |
|  | Yes | 2 | 10.5 |
| ...Tuberculosis | No | 19 | 100 |
|  | Yes | 0 | 0 |
| ...Other diseases | No | 7 | 36.8 |
|  | Yes | 12 | 63.2 |
| Had at least one complication | Unknown | 9 | 13.2 |
|  | No | 30 | 44.1 |
|  | Yes | 29 | 42.6 |
| ... Ear/Nose/Throat complications | No | 24 | 82.8 |
|  | Yes | 5 | 17.2 |
| ...Acute Otitis Media (among ENT) | No | 4 | 80 |
|  | Yes | 1 | 20 |
| ...Respiratory complications | No | 13 | 44.8 |
|  | Yes | 16 | 55.2 |
| ...Acute Respiratory Distress Syndrome (ARDS - among resp.) | Unknown | 1 | 6.2 |
|  | No | 11 | 68.8 |
|  | Yes | 4 | 25 |
| ...Pneumonia (among resp.) | Unknown | 1 | 6.2 |
|  | No | 4 | 25 |
|  | Yes | 11 | 68.8 |
| ...Pneumonia linked to COVID-19 (among pneumonia) | Unknown | 1 | 9.1 |
|  | No | 0 | 0 |
|  | Yes | 10 | 90.9 |
| ...Cardiovascular complications | No | 23 | 79.3 |
|  | Yes | 6 | 20.7 |
| ...Digestive complications | No | 22 | 75.9 |
|  | Yes | 7 | 24.1 |
| ...Liver complications | No | 22 | 75.9 |
|  | Yes | 7 | 24.1 |
| ...Renal complications | No | 26 | 89.7 |
|  | Yes | 3 | 10.3 |
| ...Neurological impairment complications | No | 25 | 86.2 |
|  | Yes | 4 | 13.8 |
| ...Osteo-articular complications | No | 29 | 100 |
|  | Yes | 0 | 0 |
| ...Thrombosis/Embolism | No | 26 | 89.7 |
|  | Yes | 3 | 10.3 |
| ...Other bacterial infection | No | 29 | 100 |
|  | Yes | 0 | 0 |
| ...Other non-bacterial infection | No | 27 | 93.1 |
|  | Yes | 2 | 6.9 |
| ...Other complications | No | 15 | 51.7 |
|  | Yes | 14 | 48.3 |
| Stayed in ICU at least once | Unknown | 8 | 11.8 |
|  | No | 54 | 79.4 |
|  | Yes | 6 | 8.8 |
| Was treated against the COVID-19 infection | Unknown | 7 | 10.3 |
|  | No | 57 | 83.8 |
|  | Yes | 4 | 5.9 |
| ...Hydroxychloroquine | No | 2 | 50 |
|  | Yes | 2 | 50 |
| ...Interferon | No | 4 | 100 |
|  | Yes | 0 | 0 |
| ...Lopinavir/Ritonavir | No | 4 | 100 |
|  | Yes | 0 | 0 |
| ...Remdesivir | No | 3 | 75 |
|  | Yes | 1 | 25 |
| ...Tenofovir | No | 4 | 100 |
|  | Yes | 0 | 0 |
| ...Ribavirin | No | 4 | 100 |
|  | Yes | 0 | 0 |
| ...Other treatment | No | 1 | 25 |
|  | Yes | 3 | 75 |
| Outcome | Deceased | 0 | 0 |
|  | Discharged and alive | 66 | 97.1 |
|  | Still hospitalised | 2 | 2.9 |

In adults, comorbidities were recorded for 2401 (67.0%) episodes, and 1884 were recorded to have more than one comorbidity. Among those, hypertension was the most frequent comorbidity, with 1481 (61.7%) occurrences, followed by chronic cardiovascular diseases (948 - 39.5%) and diabetes (660 - 27.5%). Chronic respiratory diseases were reported in 515 (21.4%) episodes, renal disease in 500 (20.8%) episodes, and neurological impairment in 386 (16.1%) episodes. Oncological pathologies (315 – 13.1%), asthma (204 – 8.5%), dementia (198 - 8.2%), liver diseases (144 - 6.0%), and immunosuppression (128 – 5.3%) were registered less often, and other comorbidities were only observed sporadically (<5%). The information about comorbidities was missing for 574 (16.0%) episodes among adults.

In children, comorbidities were recorded for 19 (27.9%) episodes and 9 were recorded to have more than one comorbidity. Respiratory diseases and asthma were the most frequent comorbidities in 4 (21.1%) episodes each, followed by haematological and oncological pathologies (3 – 15.8%), renal and liver diseases alongside neurological impairment and immunosuppression (2 – 10.5%). Finally, diabetes, hypertension, and cardio-vascular diseases were accounted in one (5.3%) episode. The information about comorbidities was missing for 7 (10.3%) episodes in children.

### Complications

Tables 2 and 3 further show lists of complications encountered in COVID-19 episodes in adults and children, respectively. Out of the 3650 episodes, 2679 (73.4%) reported at least one complication due to the COVID-19 infection, 2650 being among adults, and 29 among children. Multiple complications occurred in 1961 adult episodes and 14 children episodes. Overall 565 episodes with complications also reported a stay on Intensive Care Unit (ICU), 560 of which in adults.

In adults, respiratory complications were dominant with 2470 (93.2%) occurrences. Pneumonia was found in 2162 episodes (87.5% of respiratory complications), of which 2096 (96.9%) were found to be caused solely by COVID-19. Acute Respiratory Distress Syndrome (ARDS) was reported in 557 (22.5%) cases of respiratory complications. Renal complications were observed in 681 (25.7%) adult episodes, followed by cardiac (631 - 23.8%), liver (510 - 19.2%), digestive (498 - 18.8%), and neurological (436 - 16.5%) complications, bacterial infections other than pneumonia (329 - 12.4%). Other complications were present in less than 10% of the episodes and include thrombosis and embolisms (210 - 7.9%), ear/nose/throat complications (ENT; 193 - 7.3%; among which 8 - 4.1% - of acute otitis media), non-bacterial infections (132 – 5.0%), and osteo-articular complications (127 - 4.8%).

In children, respiratory complications were also dominant with 16 (55.2%) occurrences. Pneumonia (11 – 68.8%) was directly caused by COVID-19 in 10 (90.9%) cases of pneumonia. Those comprised ARDS in 4 (25.0%) cases in children episodes. Contrarily to adults, digestive and liver complications were the second dominant complications as they were reported in 7 (24.1%) children’s episodes with complications, followed by cardiac (6 – 20.7%), ENT (5 – 17.2%; among which one case of acute otitis media), and neurological (4 – 13.8%) complications. Finally, renal complications and thrombosis/embolism were each reported in three episodes with complications (10.3%), and non-bacterial infections in 2 (6.9%). Other bacterial infections as well as osteo-articular complications were not reported in children.

Other types of complications were registered in 1170 (43.6%) episodes with complications. Ageusia, anemia, and deconditioning syndromes were mostly reported among those. Anosmia was reported in 4 (0.2%) COVID-19 episodes. The information was missing for 596 (16.3%) of the 3650 episodes.

### Drug treatments

The distribution of drugs prescribed against the COVID-19 infection can be found in Tables 2 and 3. In 1299 (35.6% - 36.2% of episodes in adults and 5.9% of all episodes in children) episodes, one or often a combination of therapeutic agents were used. Hydroxychloroquine was the most prescribed treatment against COVID-19 (989 - 76.1%; 985 in adults and 2 in children) alongside Lopinavir/Ritonavir (675 – 52.0%; exclusively in adults). Remdesivir was the third dominant drug used to treat COVID-19 (146 – 11.2%; only once in children). Other treatments were marginally prescribed (twice for Interferon and once for Tenofovir), or not at all for Ribavirin. Note that hydroxychloroquine was almost abandoned once no more recommended by international and national authorities.

### Outcomes

Figure 3 shows the cumulative number of episodes, discharges, and deaths reported in the database. Among the 3650 episodes, 2989 (81.9%) were reported as discharged from the hospital, among which 2275 were discharged to their domicile, 230 to another hospital. 222 to a long term care facility, and 249 to other locations, such as rehabilitation centres.

**Figure 3.**
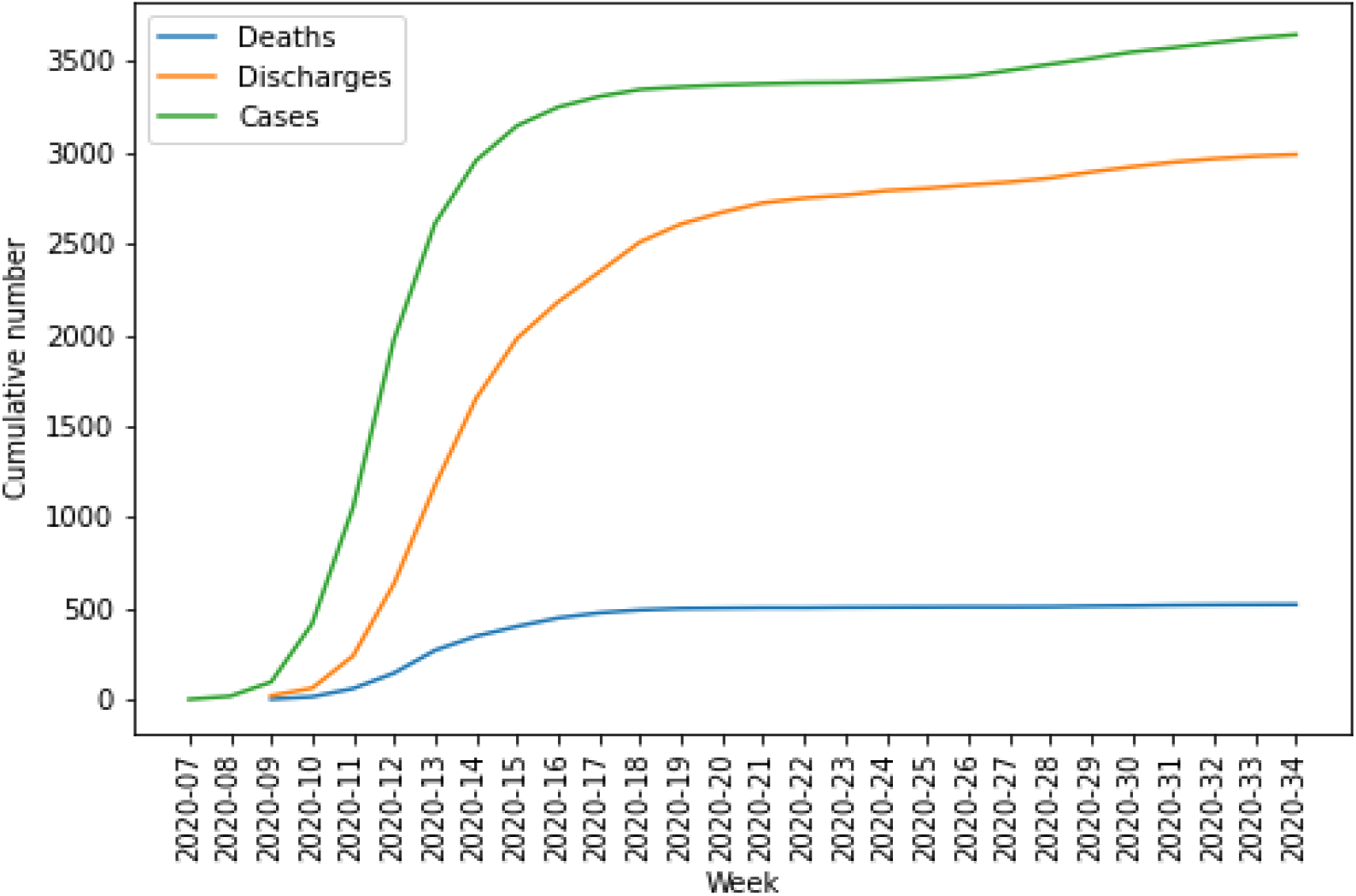
Cumulative number of episodes, discharges and deaths related to COVID-19 infection in the database.

**Figure 4.**
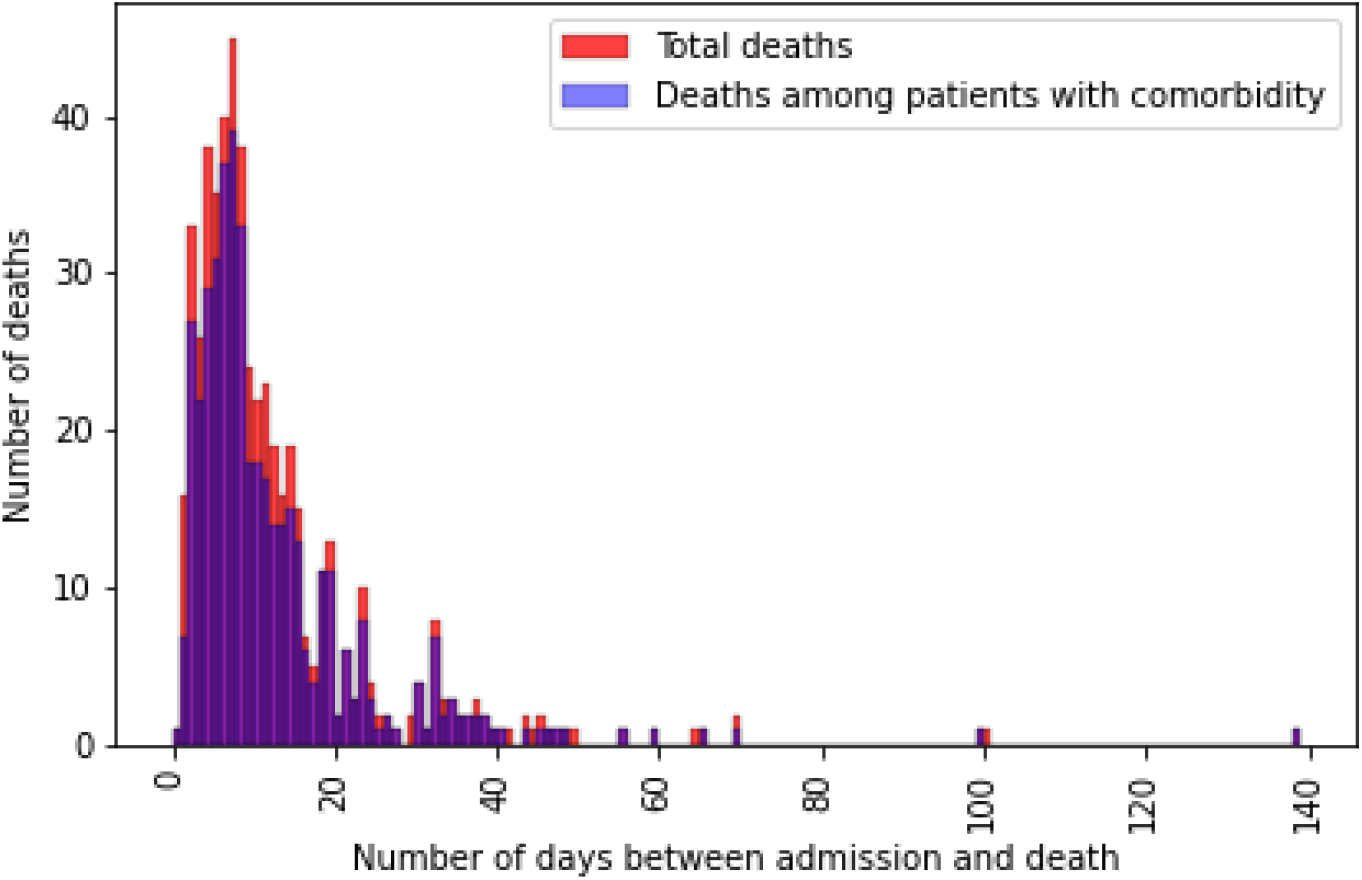
Distribution of deceased patients within the database.

**Figure 4.**
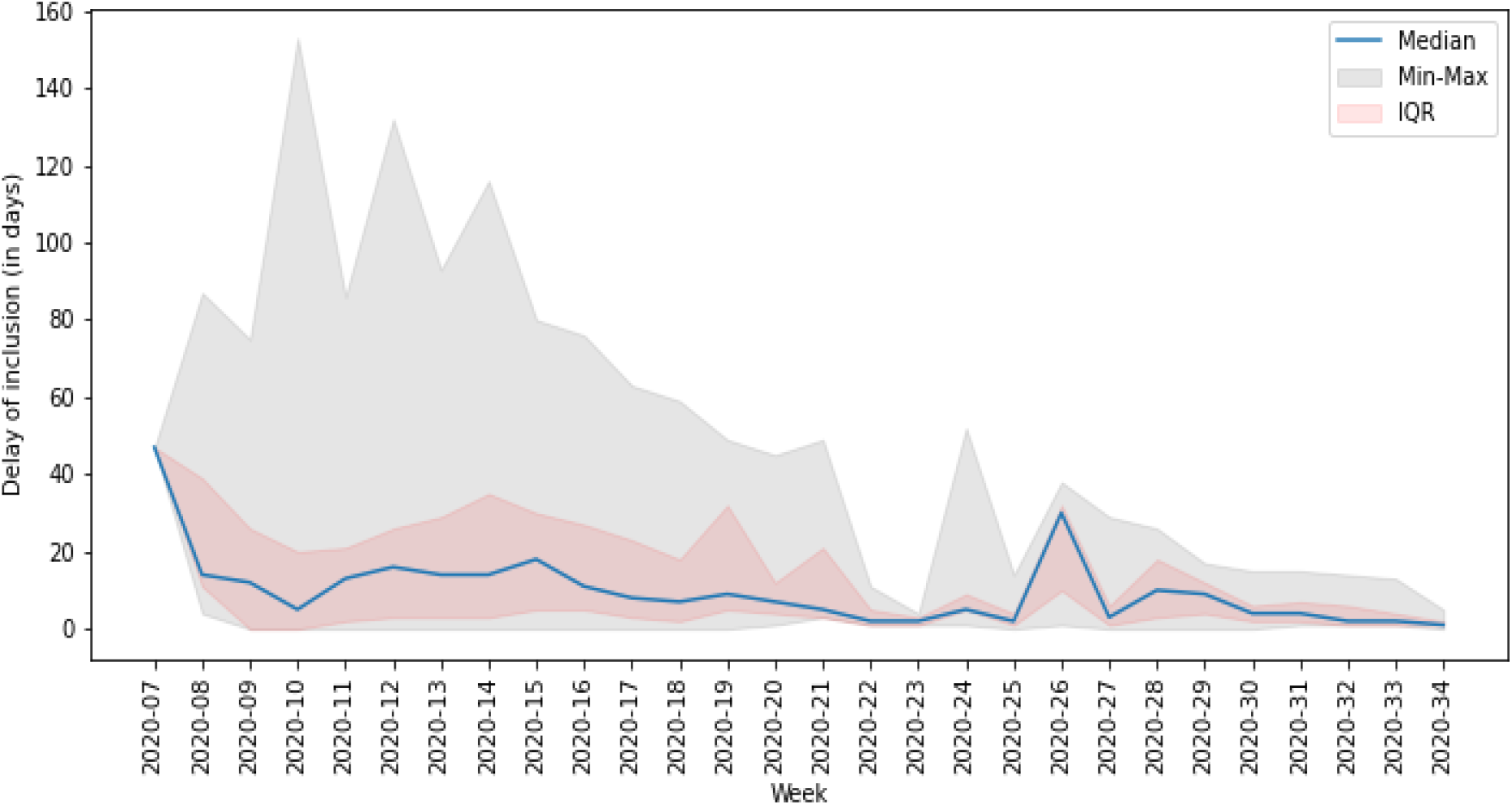
Time delay (in days) between the date of interest for the system and the inclusion of the patient in the database.

Death was reported in 527 episodes (14.4%), but none in children. The first death occurred in the week following the first recorded episode (2020-10). The number of reported deaths increased rapidly during the first 10 weeks, with only a small number of deaths occurring after week 2020-20. Figure 4 shows the number of death versus the time between the date of interest for the surveillance and the death date. Most deaths occurred in patients with comorbidities (431 – 81.8%) between the first two weeks following their diagnosis with a median time between diagnosis and death of 8 days (IQR 5-14 days). Overall, deaths has been recorded mostly in patients between 80 and 89 years (224 – 42.5%) and in patients between 70 and 79 years (147 – 27.8%) as seen in Figure 5. No deaths were reported in patients below 30.

**Figure 5.**
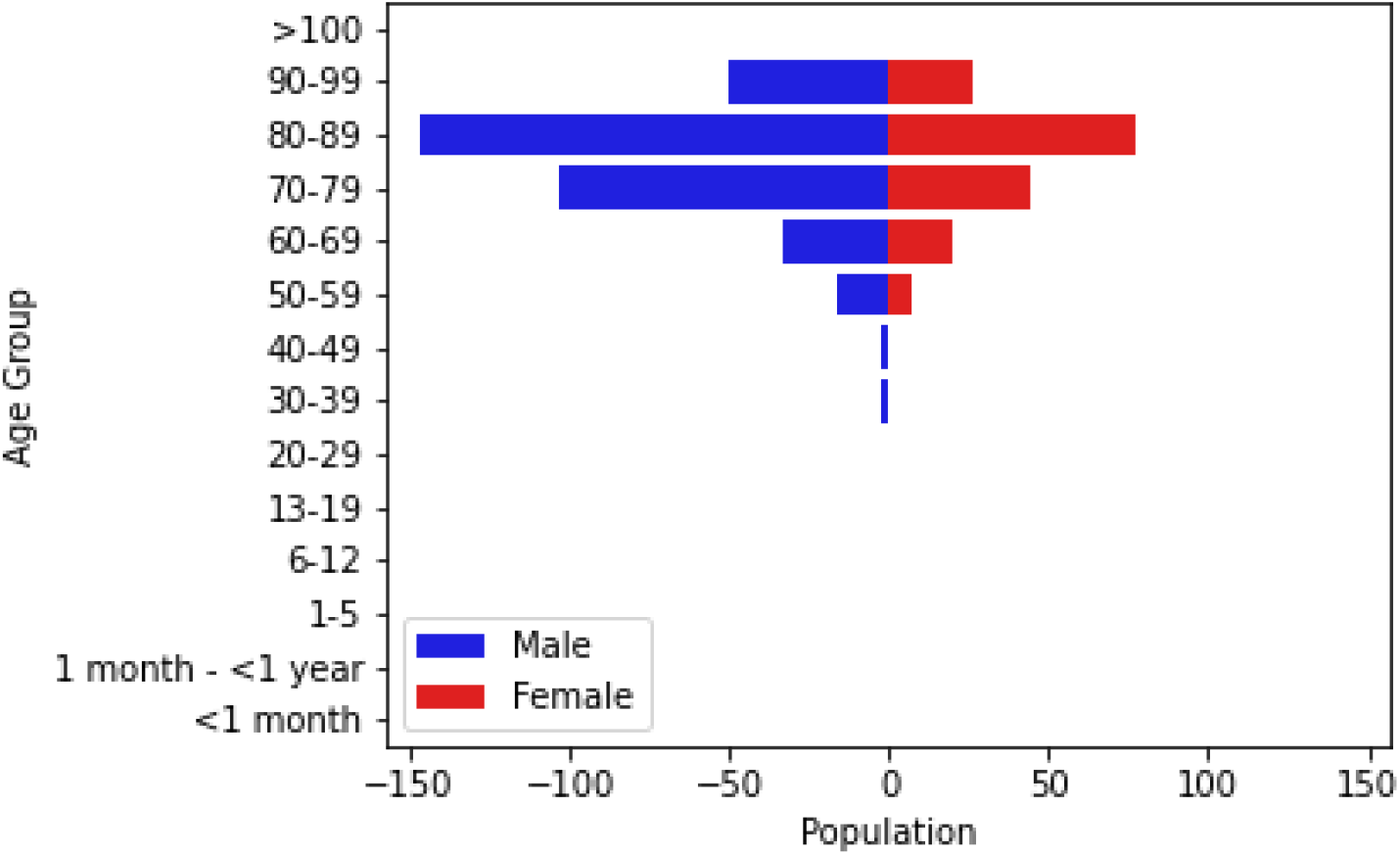
Population breakdown of the 527 deceased patients hospitalized with COVID 19 between March 1st and August 31st, 2020 by age and gender.

### Pregnant women

Pregnancy accounted for 5.9% (35) episodes of reproductive women (i.e. between 15 and 50 years old) registered in the database. Among those, 16 suffered from comorbidities (3 asthma, 3 diabetes, 2 hypertension, and 14 other comorbidities), and 14 complications occured, mostly respiratory complications in 7 episodes and ENT complications in 4. Only one pregnant woman stayed in ICU. None of them were treated against COVID-19, and no deaths were reported.

### Healthcare workers

A total of 146 (4.0%) patients were employed in a healthcare facility: one of which (0.7%) was infected in a hospital and six (4.1%) were of unknown source.

## Strengths and limitations

By using an existing surveillance system for influenza and adapting it for the surveillance of SARS-CoV-2/COVID-19 cases, we quickly developed a large harmonised dataset of hospitalised COVID-19 cases in Switzerland. The data enables assessments of disease severity, risk factors, treatments and the clinical course of the disease. The data are useful for informing public health authorities and policy makers on the impact of health regulations put in place during the pandemic and provides potential for modelling the future of the disease in the country. Due to the lack of data at the European level when the pandemic started in Europe, most models and measures taken against COVID-19 in Switzerland were based on Chinese data. The data obtained during this first wave will therefore enable healthcare workers and policy makers to respond more efficiently to the disease in the future.

Despite being rapidly set up, use of the system for real-time monitoring of the epidemic presented some challenges. Discussions with the FOPH to adapt the existing influenza database started in early February and the system was fully operational by the end of the same month, shortly after the first case was declared in Switzerland. This highlights the adaptability of the original system for surveillance of other respiratory viruses. Despite the strength of the system in place, it was difficult to obtain data within 48h of the diagnosis for most centres when faced with a high number of cases. Some hospitals started the data collection later than others, and had to enter data both retrospectively and prospectively. Other centres were overwhelmed by the large number of cases and had to focus on patient care first, while at the same time trying to train new staff who could help with data entry. The overall median time difference between the first positive laboratory test and the inclusion in the database was 13 days (IQR 3-28). Figure 4 shows the time delay throughout the pandemics. The maximum delay was observed during the height of the epidemic in Switzerland, but quickly decreased as the number of cases decreased as well. Note that other Swiss entities also requested data from several participating hospitals, making it more difficult to report episodes within the demanded 48h.

Using the same approach as the influenza database, data were checked for inconsistencies weekly and reported to the participating hospitals and the FOPH. Outliers in dates, impossible combinations of variables, length of stays and treatments, out of range numbers, and missing values were consistently checked. This allowed the participating hospitals to correct wrong and missing data rapidly. Additional checks on the data were made by the supervising physicians in each hospital based on the patients’ records to ensure medical accuracy.

By including a large number of university and cantonal hospitals, we collected 3650 cases confirmed by PCR as of September 1st, 2020 and 527 (14.4%) COVID-19 related deaths. At the same date, the mandatory reporting system put in place by the FOPH [13] registered 4405 confirmed hospitalisations related to COVID-19. As of September 3rd, 4541 confirmed hospitalisations were recorded, among which 921 (20.3%) had died during their hospital stay. 59.9% of patients were male and 40.1% female. Comorbidities were found to be hypertension in 45.9% of the patients, chronic cardiovascular diseases in 39.6%, diabetes in 20.7%, chronic respiratory diseases in 13.8%, oncological disease in 8.9%, and immunosuppression in 3.7%. Those numbers match with the proportions found in the hospital database, except for hypertensive patients, cardiovascular pathologies, and patients suffering from diabetes for which the FOPH reporting system found higher proportions. The median age of the patients in the compulsory declaration system is also slightly higher than that in our database (70 years old versus 68 years old).

We nevertheless included more than two third of the total number of hospitalisations of confirmed SARS-CoV-2/COVID-19 reported in the obligatory system of the FOPH on September 1st, 2020 (82.9%). The lower proportion of deaths in our system suggests that many patients may have died after discharge from the hospital or in another institution not captured by the system. We are also missing data on adult patients from hospitals which only agreed to contribute paediatric cases, potentially explaining the difference in the median age in both systems.

## Conclusion and outlooks

The setup of this database is a first step towards an in-depth understanding of the SARS-CoV-2/COVID-19 epidemic in Switzerland, and is a powerful tool in for the second wave, initially reported in China [14] and currently occurring in Europe. The data will allow authorities to tackle risk populations more precisely and adapt restrictions. The system was easily adapted from the influenza surveillance system in a matter of days to include and remove disease-specific information, which allowed a fast response to obtain data. However, the lack of human resources, the severity of the pandemic, and the multiplication of COVID-19 data collection in hospitals by Swiss medical societies and authorities made it difficult to obtain data rapidly. Therefore, monitoring the epidemics on time was difficult: an issue that needs to be addressed rapidly for future pandemics by working more closely with officials to insure avoiding repeated efforts.

The surveillance is still ongoing and we plan, in the future, to include patients with negative PCR results but suggestive clinical presentation combined with radiographic findings compatible with COVID-19 infection, and patients with positive SARS-CoV-2 serology. We will distinguish these 3 modes of diagnosis: while positive PCR ensures that the virus is present, CT-scans and radiology show specific features of COVID-19 only resulting in suspicions. Serology indicates that a patient was previously infected, and could therefore be of interest for hospitalisations due to complications of the infection (e.g. if the patient was not initially hospitalised).

Discussions are also ongoing for linking patients from the database to their biological samples, in order to study associations between risk factors, complications, and specific strains of the virus; and, therefore, possibly establish a microbiological surveillance of the disease.

Finally, we plan to study long-term consequences of infections since the system in place allows us to track several hospital stays in afflicted patients.

### Accessing the data

The anonymised data can be accessed through a multi-stage process. Applicants must fill a concept-sheet (supplementary material) and send it to the team in charge of the study. An Executive Committee of experts and representatives of hospital participants will review the concept. Depending on the goal of the analysis, additional ethics clearance might be needed. Data will be restricted to the request and shared through a secure platform. All steps to access the data are described in the aforementioned concept-sheet (supplementary material).

## Supporting information

Supplementary File 1 - Case Report Form

Supplementary File 2 - Concept Sheet for data acess

## Acknowledgments

The authors would like to thank all the participating centres’ team, study nurses, and physicians for their hard work and commitment to the study.

This work was supported by the Swiss Federal Office of Public Health under reference 333.0-20/1. OK acknowledges additional support from the Swiss National Science Foundation (SNF) via grant #163878.

PWS has received grant support from the Career funding program “Filling the Gap” of the Medical Faculty of the University of Zurich With contributions of the Clinical Research Center, Geneva University Hospitals and Faculty of Medicine, Geneva.

This study was submitted and approved by the Geneva Ethics Committee (CCER) and by all hospitals’ local Ethics Committee through the Swissethics BASEC submission system, under reference 2020-00827.

## Contributions

AT has set up the system, including coding the database, checking for data inconsistencies, and written the scripts for analyses. He also drafted an corrected the present manuscript.

AI has helped setting the first version of the CRF, provided medical expertise, helped drafting the first version of the manuscript. She is also the project leader for the Hôpitaux Universitaire de Genève.

CB, LS, NT, AW, DF, PWS, MV, LD, MB, DVG, CK, AC, TR, YN, RG, UH, CB, FZ, SBS, NC, PZ, AU, and ANL were part of the data collection teams as local project leaders, provided feedback on the CRF and on each iteration of the manuscript.

CG lead the project for the Federal Office of Public Health. She helped designing the CRF from an official perspective and provided administrative support.

MR helped designing the CRF and assessing data quality throughout the data collection period. OK provided feedback and correction for each iteration of the manuscript.

