## Supplementary File 1 - Case Report Form for "SARS-CoV-2/COVID-19 hospitalised patients in Switzerland: a prospective cohort profile"

### Inclusion

Record ID (patient identifier) __________________________________

**Hospital based surveillance of COVID-19 cases in Switzerland**

**Patient-level information**

**Each new record is a distinct COVID-19 Episode related to a patient.**
**In case a specific patient undergoes more than one episode, please create a new record to report each additional episode.**

Is this another COVID-19 episode from a same patient ?

◯ No (this is the patient's first episode)
◯ Yes (the first episode record has been already reported)
◯ Still to be confirmed

ID of first episode of this patient

ID is the number on the right of the full CenterID-ID identifier, e.g. 123-456

__________________________________
([0-9999])

Center (or consortium) where the first episode was created

◯ CHUV (Lausanne) ◯ EOC (Lugano) ◯ HFR (Fribourg) ◯ Hirslanden AG ZH (Zurich) ◯ Hopital VS (Sion) ◯ HUG (Geneva) ◯ Inselspital (Bern) ◯ KISPI (Basel) ◯ KISPI (Zurich) ◯ KSA (Aarau) ◯ KSGR (Graubuenden) ◯ KSNW (Niedwalden) ◯ KSSG (St.Gallen) & consortium ◯ KSW (Winterthur) ◯ LUKS (Luzern) ◯ OKS (St.Gallen) ◯ Spitaeler SH (Schaffhausen) ◯ STGAG KSM (Muensterlingen) ◯ USB (Basel) ◯ USZ (Zurich)
(your current center: [user-dag-label])

#### Checking inclusion criteria

| | |
|---|---|
| Hospitalised for more than 24 hours | ◯ Yes ◯ No |
| Laboratory-confirmed COVID-19 diagnosis | ◯ Yes ◯ No |

#### Patient's inclusion

| | |
|---|---|
| Confirm inclusion ? | ☐ Yes (include patient) |
| Inclusion date | ________________________________ |
| ID of user checking the inclusion | ________________________________ |
| Current date | ◯ 2020-10-29<br>((last date form is saved)) |

### Demography

#### Demography

| | |
|---|---|
| Year of birth | ________________________ |
| Is the patient 6 years old or less ? | ◯ No<br>◯ Yes |
| Birth Month | ________________________ |
| Gender | ◯ Male ◯ Female ◯ Other |

Height, Weight and BMI will be evaluated during each individual hospitalisation event

### Case Declaration

Starting date of COVID-19 symptoms ______________________

#### Exposure factors

Type of exposure
- ◯ Community acquired
- ◯ Nosocomial (> 5 days)
- ◯ Unknown

Where was the patient contaminated?
- ◯ Household
- ◯ School / Kindergarten /Daycare
- ◯ Unknown

Employed in a healthcare facility ?
- ◯ No
- ◯ Yes
- ◯ Unknown

Employed in a microbiology laboratory?
- ◯ No
- ◯ Yes
- ◯ Unknown

#### Sample

Date and time of lab sample collection ______________________

...check if date/time of sample may NOT be exact
- ◯ exact date and time
- ◯ exact date / estimated time
- ◯ estimated date and time

(optional)

Type of sample
- ◯ Nasal swab
- ◯ Throat swab
- ◯ Nasopharyngeal swab
- ◯ Tracheal aspiration
- ◯ Broncho-alveolar lavage
- ◯ Other...

...please, specify sample type ______________________

In which service was the sample taken ?
- ◯ Medicine
- ◯ Geriatrics
- ◯ Intensive Care
- ◯ Surgery
- ◯ Paediatrics
- ◯ Emergency Room
- ◯ Other...

...please, specify where the sample was taken ______________________

| | |
|---|---|
| Laboratory confirmation method | ◯ RT-PCR<br>◯ Other... |
| ...please, specify confirmation method | __________________________________ |
| Was there another sample taken for laboratory testing? | ◯ No<br>◯ Yes |

#### Second sample

| | |
|---|---|
| Date and time of lab sample collection | __________________________________ |
| ...check if date/time of sample may NOT be exact | ◯ exact date and time<br>◯ exact date / estimated time<br>◯ estimated date and time<br>(optional) |
| Type of sample | ◯ Nasal swab<br>◯ Throat swab<br>◯ Nasopharyngeal swab<br>◯ Tracheal aspiration<br>◯ Broncho-alveolar lavage<br>◯ Other... |
| ...please, specify sample type | __________________________________ |
| In which service was the sample taken ? | ◯ Medicine<br>◯ Geriatrics<br>◯ Intensive Care<br>◯ Surgery<br>◯ Paediatrics<br>◯ Emergency Room<br>◯ Other... |
| ...please, specify where the sample was taken | __________________________________ |
| Laboratory confirmation method | ◯ RT-PCR<br>◯ other... |
| ...please, specify confirmation method | __________________________________ |
| Laboratory test result | ◯ Positive<br>◯ Negative |
| Was there another sample taken for laboratory testing? | ◯ No<br>◯ Yes |

#### Third sample

| | |
|---|---|
| Date and time of lab sample collection | __________________________________ |
| ...check if date/time of sample may NOT be exact | ◯ exact date and time<br>◯ exact date / estimated time<br>◯ estimated date and time<br>(optional) |
| Type of sample | ◯ Nasal swab<br>◯ Throat swab<br>◯ Nasopharyngeal swab<br>◯ Tracheal aspiration<br>◯ Broncho-alveolar lavage<br>◯ Other... |
| ...please, specify sample type | __________________________________ |
| In which service was the sample taken ? | ◯ Medicine<br>◯ Geriatrics<br>◯ Intensive Care<br>◯ Surgery<br>◯ Paediatrics<br>◯ Emergency Room<br>◯ Other... |
| ...please, specify where the sample was taken | __________________________________ |
| Laboratory confirmation method | ◯ RT-PCR<br>◯ Other... |
| ...please, specify confirmation method | __________________________________ |
| Laboratory test result | ◯ Positive<br>◯ Negative |
| Was there another sample taken for laboratory testing? | ◯ No<br>◯ Yes |

#### Fourth sample

| | |
|---|---|
| Date and time of lab sample collection | __________________________________ |
| ...check if date/time of sample may NOT be exact | ◯ exact date and time<br>◯ exact date / estimated time<br>◯ estimated date and time<br>(optional) |

| | |
|---|---|
| Type of sample | ◯ Nasal swab<br>◯ Throat swab<br>◯ Nasopharyngeal swab<br>◯ Tracheal aspiration<br>◯ Broncho-alveolar lavage<br>◯ Other... |
| ...please, specify sample type | __________________________________ |
| In which service was the sample taken ? | ◯ Medicine<br>◯ Geriatrics<br>◯ Intensive Care<br>◯ Surgery<br>◯ Paediatrics<br>◯ Emergency Room<br>◯ Other... |
| ...please, specify where the sample was taken | __________________________________ |
| Laboratory confirmation method | ◯ RT-PCR<br>◯ Other... |
| ...please, specify confirmation method | __________________________________ |
| Laboratory test result | ◯ Positive<br>◯ Negative |

### Admission

| | |
|---|---|
| Please confirm that the patient is rehospitalised following complications of this same COVID-19episode! | ☐ same COVID-19 episode |
| Entry date in the hospital | __________________________________ |

#### Patient's admission

| | |
|---|---|
| Where was the patient hospitalised ? | ◯ Medicine<br>◯ Geriatrics<br>◯ Intensive Care<br>◯ Surgery<br>◯ Paediatrics<br>◯ Emergency Room<br>◯ Other... |
| ...please, specify where he/she has been diagnosed | __________________________________ |
| Was the patient hospitalised in an unit dedicated to COVID-19) | ◯ No<br>◯ Yes<br>◯ Unknown |
| [Only applicable for hospital consortia]<br><br>If admission didn't occur in the main hospital of your consortium, please provide the ID of the subsidiary hospital in your consortium | ◯ 2<br>◯ 3<br>◯ 4<br>◯ 5<br>◯ 6<br>◯ 7<br>◯ 8<br>◯ 9<br>((optional)) |
| ...code of Unit/Building | __________________________________<br>(optional) |
| Origin (pre-hospitalisation) | ◯ Domicile<br>◯ Long term care<br>◯ Other hospital<br>◯ Other... |
| ...please, specify origin | __________________________________ |
| Was the patient in contact with a healthcare personnel prior to hospitalisation (by phone or consultation) ? | ◯ Yes<br>◯ No<br>◯ Unknown |

#### Height and Weight during hospitalisation

Height

previously reported height (if applicable):
[height][previous-instance]

______________________
([cm])

Weight

______________________
([kg])

BMI

______________________
([kg/m2])

Obesity

○ No ○ Yes ○ Unknown

This is only a warning message:
the BMI calculation and obesity status do not match.
Please check the given values.

Note that the WHO classification based on BMI is lacking subtleties, so this warning is only present to raise awareness on a possible error. It does not imply that there is indeed an error.

#### Symptoms

Severity (CURB-65 score)

- ☐ Confusion (abbreviated Mental Test Score < 9)
- ☐ Urea (BUN > 19 mg/dL or 7 mmol/L)
- ☐ Respiratory rate > 30 per minute
- ☐ Blood pressure: diastolic < 60 or systeolic < 90 mmHg
- ☐ Age >= 65 years
- ☐ None of the above

Severity (for children)

- ☐ Respiratory distress
- ☐ Oxygen saturation < 92%
- ☐ Evidence of severe clinical dehydration or clinical shock
- ☐ Altered conscious level
- ☐ None of the above

Total score (each choice counts for 1)

______________________
(0-1 points: low risk >1 points: high risk)

Additional symptoms

- ☐ Cough
- ☐ Rhinitis
- ☐ Diarrhoea
- ☐ Fever
- ☐ None of the above

What was the highest temperature recorded?

______________________
(in degrees celsius)

### Clinical Complementary Information

#### Co-morbidities

Usual good health (no co-morbidities)

◯ No ◯ Yes

| | |
|---|---|
| Chronic respiratory disease | ◯ No ◯ Yes ◯ Unknown |
| ... please specify | __________________ |
| Asthma | ◯ No ◯ Yes ◯ Unknown |
| Diabetes | ◯ No ◯ Yes ◯ Unknown |
| Hypertension | ◯ No ◯ Yes ◯ Unknown |
| Chronic cardiovascular disease | ◯ No ◯ Yes ◯ Unknown |
| Chronic renal disease | ◯ No ◯ Yes ◯ Unknown |
| Chronic liver disease | ◯ No ◯ Yes ◯ Unknown |
| Chronic neurological impairment | ◯ No ◯ Yes ◯ Unknown |
| Hematological pathology with immuno-suppression | ◯ No ◯ Yes ◯ Unknown |
| Oncological pathologies | ◯ No ◯ Yes ◯ Unknown |
| Rheumatological pathology with immuno-suppression | ◯ No ◯ Yes ◯ Unknown |
| Dementia | ◯ No ◯ Yes ◯ Unknown |
| Transplant (solid organs) | ◯ No ◯ Yes ◯ Unknown |
| HIV-positive | ◯ No ◯ Yes ◯ Unknown |
| Immuno-suppressive treatment | ◯ No ◯ Yes ◯ Unknown |
| Tuberculosis | ◯ No ◯ Yes ◯ Unknown |
| Others | ◯ No ◯ Yes ◯ Unknown |
| .... please specify | __________________ |

#### Other risk factors

| | |
|---|---|
| Pregnancy | ◯ No ◯ Yes ◯ Unknown |
| Postpartum < 4 weeks | ◯ No ◯ Yes ◯ Unknown<br>(Women who gave birth in the 4 weeks before the COVID-19 episode) |
| Premature < 24 months | ◯ No ◯ Yes ◯ Unknown<br>(Premature children aged < 24 months) |
| ...please specify the gestational week the child was born in | ________________________________<br>(Number between 0 and 38) |
| ...please specify weight at birth | ________________________________<br>(in kg) |
| Smoking | ◯ No ◯ Yes ◯ Unknown |
| Is the patient under an ACE inhibitor? | ◯ No ◯ Yes ◯ Unknown |
| Was the patient prescribed or treated with cardiovascular medications (during or prior to hospitalisation)? | ◯ No ◯ Yes ◯ Unknown |
| Charlson Comorbidity Index (CCI)<br><br>[mdcalc Calculator]<br><br>Charlson M, Szatrowski TP, Peterson J, Gold J. Validation of a combined comorbidity index. J Clin Epidemiol. 1994;47(11):1245-51. PMID: 7722560 | ________________________________ |

#### Antiviral treatment (against COVID-19)

| | |
|---|---|
| Prophylactic treatment (against COVID-19) | ◯ No ◯ Yes ◯ Unknown |
| Treatment of confirmed infection (against COVID-19) | ◯ No ◯ Yes ◯ Unknown |
| Name of the treatment | ☐ Chloroquin ☐ Interferon<br>☐ Lopinavir/Ritonavir ☐ Remdesivir<br>☐ Tenofovir ☐ Ribavirin<br>☐ Other... |
| ...please, specify (name of treatment) | ________________________________ |
| Starting date of the treatment | ________________________________<br>((if available)) |

Ending date of the treatment

__________________________
((if available))

#### Stay in Intermediate care

Did the patient stay in intermediate care ?

◯ No ◯ Yes ◯ Unknown

#### Intermediate care (first stay)

Intermediate care entry date

__________________________
((if available))

Intermediate care exit date

__________________________
((if available))

Non-invasive ventilation ◯ No ◯ Yes ◯ Unknown

Any additional stay in intermediate care to report ? ◯ No ◯ Yes

#### Intermediate care (second stay)

Intermediate care entry date

__________________________
((if available))

Intermediate care exit date

__________________________
((if available))

Non-invasive ventilation ◯ No ◯ Yes ◯ Unknown

Any additional stay in intermediate care to report ? ◯ No ◯ Yes

#### Intermediate care (third stay)

Intermediate care entry date

__________________________
((if available))

Intermediate care exit date

__________________________
((if available))

Non-invasive ventilation ◯ No ◯ Yes ◯ Unknown

#### Stay in Intensive care

Did the patient stay in intensive care ?

◯ No ◯ Yes ◯ Unknown

#### Intensive care (first stay)

Intensive care entry date

__________________________
((if available))

Intensive care exit date

__________________________
((if available))

Non-invasive ventilation ◯ No ◯ Yes ◯ Unknown

Invasive ventilation ◯ No ◯ Yes ◯ Unknown

Extra-Corporeal Membrane Oxygenation (ECMO) ◯ No ◯ Yes ◯ Unknown

Any additional stay in intensive care to report ? ◯ No ◯ Yes

#### Intensive care (second stay)

Intensive care entry date

__________________________
((if available))

Intensive care exit date

__________________________
((if available))

Non-invasive ventilation ◯ No ◯ Yes ◯ Unknown

Invasive ventilation ◯ No ◯ Yes ◯ Unknown

Extra-Corporeal Membrane Oxygenation (ECMO) ◯ No ◯ Yes ◯ Unknown

Any additional stay in intensive care to report ? ◯ No ◯ Yes

#### Intensive care (third stay)

| | |
|---|---|
| Intensive care entry date | ________________ ((if available)) |
| Intensive care exit date | ________________ ((if available)) |
| Non-invasive ventilation | ◯ No ◯ Yes ◯ Unknown |
| Invasive ventilation | ◯ No ◯ Yes ◯ Unknown |
| Extra-Corporeal Membrane Oxygenation (ECMO) | ◯ No ◯ Yes ◯ Unknown |

#### Complications (probably related to COVID-19)

Did the patient have any complications ?

◯ No ◯ Yes ◯ Unknown

| | |
|---|---|
| Ear/Nose/Throat (ENT) diseases | ◯ No ◯ Yes ◯ Unknown |
| Acute Otitis Media | ◯ No ◯ Yes ◯ Unknown |
| Respiratory diseases | ◯ No ◯ Yes ◯ Unknown |
| Acute respiratory distress syndrome | ◯ No ◯ Yes ◯ Unknown |
| Pneumonia | ◯ No ◯ Yes ◯ Unknown |
| ...pneumonia code [see pneumonia classification] | ◯ PN1 ◯ PN2 ◯ PN3 ◯ PN4 ◯ PN5 ◯ Lobar pneumonia ◯ Other |
| ...was the pneumonia associated with COVID-19? | ◯ No ◯ Yes ◯ Unknown |
| Cardiac disease | ◯ No ◯ Yes ◯ Unknown |
| Digestive disease | ◯ No ◯ Yes ◯ Unknown |
| Liver disease | ◯ No ◯ Yes ◯ Unknown |
| Renal disease | ◯ No ◯ Yes ◯ Unknown |
| Neurological impairment | ◯ No ◯ Yes ◯ Unknown |

| | |
|---|---|
| Osteo-articular disease | ◯ No ◯ Yes ◯ Unknown |
| Thrombosis/Embolism | ◯ No ◯ Yes ◯ Unknown |
| Other bacterial infections (excepted pneumonia) | ◯ No ◯ Yes ◯ Unknown |
| Other non-bacterial infections | ◯ No ◯ Yes ◯ Unknown |
| Other complications... | ◯ No ◯ Yes |
| ...please, specify (complications) | ______________________ |

#### Antibiotic treatment (against complications)

| | |
|---|---|
| Antibiotic treatment taken (against complications) | ◯ No ◯ Yes ◯ Unknown |
| Code of given antibiotics (main)<br>[see list of AB codes] | ______________________<br>([code required - 0 if n/a]) |
| Code of given antibiotics (additional) | ______________________<br>([optional]) |
| Code of given antibiotics (additional) | ______________________<br>([optional]) |
| Code of given antibiotics (additional) | ______________________<br>([optional]) |
| Code of given antibiotics (additional) | ______________________<br>([optional]) |
| Antifungal treatment taken (against complications) | ◯ No ◯ Yes ◯ Unknown |
| Cortico-steroids treatment taken (against complications) | ◯ No ◯ Yes ◯ Unknown |

### Patient Follow Up

#### Transfers

Was the patient transferred during hospitalisation? ◯ Yes ◯ No ◯ Unknown

... the patient was transferred in
- ☐ Medicine
- ☐ Geriatrics
- ☐ Intensive Care
- ☐ Surgery
- ☐ Paediatrics
- ☐ Others...

... please specify (one item only) ______________________

#### Patient's destination

Deceased ◯ Yes ◯ No ◯ Unknown

... death occured
- ◯ during hospitalisation
- ◯ after being discharged

Date of death ______________________

Was the death caused by COVID-19? ◯ No ◯ Yes ◯ Unknown

Destination
- ◯ Domicile
- ◯ LTC Facility
- ◯ Another hospital
- ◯ Other
- ◯ Unknown

... please specify destination ______________________

Was the patient transferred to an hospital participating to this surveillance system?
- ◯ No
- ◯ Yes

| | |
|---|---|
| In which participating hospital was the patient transferred?<br><br>Make sure you give the patient ID to the hospital he/she is being transferred to in order to ease the follow-up process! | ◯ CHUV (Lausanne)<br>◯ EOC (Lugano)<br>◯ HFR (Fribourg)<br>◯ Hirslanden AG ZH (Zurich)<br>◯ Hopital VS (Sion)<br>◯ HUG (Geneva)<br>◯ Inselspital (Bern)<br>◯ KISPI (Basel)<br>◯ KISPI (Zurich)<br>◯ KSA (Aarau)<br>◯ KSGR (Graubuenden)<br>◯ KSNW (Niedwalden)<br>◯ KSSG (St.Gallen) & consortium<br>◯ KSW (Winterthur)<br>◯ LUKS (Luzern)<br>◯ OKS (St.Gallen)<br>◯ Spitaeler SH (Schaffhausen)<br>◯ STGAG KSM (Muensterlingen)<br>◯ USB (Basel)<br>◯ USZ (Zurich) |
| Why was the patient transferred to another hospital? | ◯ Lack of space<br>◯ Favourable evolution (the patient was put in recovery care)<br>◯ Unfavourable evolution (the patient needed to be put in intensive care)<br>◯ Unknown |
| Discharging date from hospital | ______________________________ |
| Did the patient leave with any sequelae requiring post-discharge treatment? | ◯ Yes ◯ No ◯ Unknown |

#### Comments

| | |
|---|---|
| Comments | ______________________________ |
