## Supplementary File 2 - Concept Sheet for data acess for "SARS-CoV-2/COVID-19 hospitalised patients in Switzerland: a prospective cohort profile"

**CONCEPT SHEET: MULTI-CENTRIC ANALYSIS**

**NOTE: Any analysis, which is not done for epidemiological surveillance purpose only, needs to be submitted for additional ethics approval through an amendment or a new submission**

| **Database of interest:** | ☐ COVID-19/SARS-CoV-2  ☐ Influenza |
| --- | --- |
| **Other sources of data** | ☐ Yes (specify)  ☐ No |
| **Title:** |  |
| **Concept Lead:** | *(please provide name, email address, address, and phone number)* |
| **Collaborators:** | *(please provide names, email addresses, and roles in the analysis)* |
| **Data Manager:** | *(please provide name, email address, address, and phone number)* |
| **Lead Statistician:** | *(please provide name, email address, address, and phone number)* |
| **Additional participants:** | *(please provide names, email addresses, and roles in the analysis)* |
| **Where will statistical analyses be done?** |  |
| **Abstract:** (±200 words) | Background and objectives  Methods |
| **Project outline:**  (±1000 words) | Background  Objectives and hypotheses  Study design  Eligibility criteria  Key variables and definitions  Outcomes  References |
| **Ethics:** | ☐ This concept fulfils a need in epidemiological surveillance and does not require additional ethics approval.  ☐ This concept uses the data for other purposes and, therefore, requires additional ethics approval through an amendment.  ☐ This concept adds other sources of data. This requires additional ethics approval through a new multi-centric submission.  *Describe*: |
| **Dataset:** | Please provide a list of variables needed for the analysis, based on the Codebooks available on the corresponding website:   - COVID-19: <https://www.unige.ch/medecine/hospital-covid/> - Influenza: <https://www.unige.ch/medecine/hospital-flu/> |
| **Target journal(s):** |  |
| **Milestones:** | Circulation of concept sheet: <date>  Circulation of draft paper: <date>  Submission to target journal: <date> |

By submitting this concept, the team and the leader understand that the dataset will be limited to the provided list of variables and shared on a secure platform (Renku - [renkulab.io](https://renkulab.io/)). Additional analyses not described in the concept proposal need separate approval. Any data sharing to third parties or to persons who do not participate in the analysis is strictly forbidden as per the terms of the original protocol and study agreements.

The latest forms (e.g. description of procedure, variable description, protocol, concept sheet template) can be found on the corresponding websites.

**Please read the documents** **carefully before submitting a concept proposal**.

**Accepted abstracts will be made publically available on those websites to avoid duplication of efforts**.

☐ I have read and agreed with the terms and conditions described above

Date & place:

Name & signature:

**Next Steps**

Thank you for preparing a concept proposal for analysis of hospital-sentinel data. Here are the steps for submitting your concept:

1. Before submitting the concept sheet, please **ensure all sections have been completed** or marked not applicable, the document is clean (all edits and comments are removed), and references have been added.
2. Once the document is ready for circulation to the Executive Committee, **send it to the following contacts**:
   a. For the COVID-19 Database
      i. Dr Amaury Thiabaud –
      ii. Dr Maroussia Roelens –
      iii. Prof Olivia Keiser –
   b. For the Influenza Database
      i. Dr Amaury Thiabaud –
      ii. Mr Erol Orel –
      iii. Prof Olivia Keiser –

The Institute of Global Health team will review the concept prior to circulation to the Executive Committee. This Committee has 14 days to review the concept sheet, as agreed in the study agreements. If you have questions about the form content, please contact one of the persons listed above.
